# Antibody Responses to SARS-CoV-2 after Infection or Vaccination in Children and Young Adults with Inflammatory Bowel Disease

**DOI:** 10.1101/2021.06.12.21258810

**Authors:** Joelynn Dailey, Lina Kozhaya, Mikail Dogan, Dena Hopkins, Blaine Lapin, Katherine Herbst, Michael Brimacombe, Kristen Grandonico, Faith Karabacak, John Schreiber, Bruce Tsan-Liang Liang, Juan C. Salazar, Derya Unutmaz, Jeffrey S. Hyams

**Author notes:** Correspondence: Jeffrey S. Hyams, MD, Division of Digestive Diseases, Hepatology, and Nutrition, Connecticut Children’s Medical Center, 282 Washington Street, Hartford, CT, 06106; Derya Unutmaz, MD, Jackson Laboratory for Genomic Medicine, 10 Discovery Drive, Farmington, CT, 06032. These authors contributed equally (Joelynn Dailey, Lina Kozhaya, Mikail Dogan). Shared senior authorship (Derya Unutmaz, Jeffrey S. Hyams).

## Abstract

**Background:** Characterization of neutralization antibodies to SARS-CoV-2 infection or vaccination in children and young adults with inflammatory bowel disease (IBD) receiving biologic therapies is crucial.

**Methods:** **W**e performed a prospective longitudinal cohort study evaluating SARS-CoV-2 Spike protein receptor binding domain (S-RBD) IgG positivity along with consistent clinical symptoms in patients with IBD receiving infliximab or vedolizumab. Serum was also obtained following immunization with approved vaccines. IgG antibody to the spike protein binding domain of SARS-CoV-2 was assayed with a fluorescent bead-based immunoassay that takes advantage of the high dynamic range of fluorescent molecules using flow cytometry. A sensitive and high-throughput neutralization assay that incorporates SARS-CoV-2 Spike protein onto a lentivirus and measures pseudoviral entry into ACE2 expressing HEK-293 cells was used.

**Results:** 436 patients were enrolled (mean age 17 years, range 2-26 years, 58% male, 71% Crohn’s disease, 29% ulcerative colitis, IBD-unspecified). 44 (10%) of enrolled subjects had SARS-CoV-2 S-RBD IgG antibodies. Compared to non-IBD adults (ambulatory) and hospitalized pediatric patients with PCR documented SARS-CoV-2 infection, S-RBD IgG antibody levels were significantly lower in the IBD cohort and by 6 months post infection most patients lacked neutralizing antibody. Following vaccination (n=33) patients had a 15-fold higher S-RBD antibody response in comparison to natural infection, and all developed neutralizing antibodies to both wild type and variant SARS-CoV-2.

**Conclusions and Relevance:** The lower and less durable SARS-CoV-2 S-RBD IgG response to natural infection in IBD patients receiving biologics puts them at risk of reinfection. The robust response to immunization is likely protective.

**Summary:** Our study showed a low and poorly durable SARS-CoV-2 S-RBD neutralizing IgG response to natural infection in IBD patients receiving biologics potentially putting them at risk of reinfection. However, they also had a robust response to immunization that is likely protective

## Introduction

Individuals with inflammatory bowel disease (IBD) potentially have higher risk of symptomatic or severe SARS-CoV-2 disease because of the immunosuppressive therapies they receive. However, results from large multicenter international studies performed in adults and children with IBD to date show similar severity of illness and risk of hospitalization or death compared with age-matched individuals without IBD (1-3). Nonetheless, corticosteroid therapy, thiopurine therapy, or combination anti-TNF/thiopurine at the time of infection have been associated with more severe disease (1, 4).

SARS-CoV-2 elicits a robust humoral and cellular immune response (5-12). A strong antibody response generated by effector B cells is critical for the eventual clearance of viruses, and the development of potent neutralizing antibodies is a major part of the memory response that prevents reinfection. Indeed, SARS-CoV-2 elicits virus-specific IgM, IgG and IgA, and neutralizing IgG antibodies (nAbs) following 7-14 days post-infection (9, 10). IgG levels to SARS-CoV-2 nucleocapsid (N) and spike proteins increase gradually following infection. Antibodies that bind to the receptor binding domain (RBD) of spike protein may neutralize viral entry into cells and play an important role in the protective immune response to SARS-CoV-2 (11). A recent study from the United Kingdom of 7000 adult patients receiving either the biological therapies infliximab (anti-TNFα) or vedolizumab (anti-α4β7 integrin) (13) revealed lower seroprevalence of anti-nucleocapsid SARS-CoV-2 in infliximab-treated compared to vedolizumab-treated patients (3.4% (161/4685) vs 6.0% (134/2250), p<0.0001) though rates of symptomatic and polymerase chain reaction (PCR) proven SARS-CoV2 infection were the same in both groups. In patients with confirmed SARS-CoV-2 infection, seroconversion was observed in fewer infliximab-treated than vedolizumab-treated patients (48% (39/81) vs 83% (30/36), p=0.00044) and the magnitude of anti-SARS-CoV-2 reactivity was lower.

In the late winter/spring 2020 Connecticut (U.S.A) was a “hot spot” for SARS-CoV-2 infection presenting a unique opportunity to perform longitudinal surveillance on a geographically defined population of children and young adults with IBD receiving biologics in our ambulatory infusion center. Here we report IgG S-RBD antibody response to SARS-CoV-2 infection and its durability over time, assess the neutralization capability of serum from our patient cohort to native and variant virus, and the relationship of this humoral response to clinical expression of COVID-19. The recent availability of SARS-CoV2 vaccine for young individuals further allowed us to assess SARS-CoV2 S-RBD IgG responses following vaccination.

## Materials and Methods

### Subjects

Between May 2020 and April 2021, we performed a single-center prospective longitudinal study evaluating seropositivity to SARS-CoV2 spike protein binding domain in an ambulatory infusion center in Farmington, CT. Patient eligibility included: (1) diagnosis of IBD, (2) ≥3 infusions with either infliximab or vedolizumab prior to study entry, and (3) assent/consent. At each serum sampling (approximately every 8-16 weeks), participants/parents completed a paper questionnaire capturing clinical information suggestive of clinical signs or symptoms of COVID-19 in the study participant or household members, and whether there was PCR documentation of SARS-CoV-2 infection dating back to January 2020 in the participant or a family member. Demographic characteristics, IBD activity, and concomitant medications were recorded. Data were entered into a Research Electronic Data Capture (REDCap) system (14).

Serum for antibody titers was obtained at the time of the biologic infusion. Approximately 5 ml of blood was drawn in a serum separator collection tube. Tubes were spun and stored at 4°C for ≤2 days until transport to Jackson Laboratories for Genomic Medicine (JAX-GM), where serum was extracted and stored at -80°C. Serum was obtained from non-IBD pediatric patients hospitalized with SARS-CoV-2 infection (n=11). Serum from adults without IBD (UConn Healthcare workers, n=23) with SARS-CoV-2 infection who remained ambulatory were collected at the time of diagnosis (baseline) and 2, 4, 6 months. Consent and assent were obtained from the participant/family in all cases. This study was reviewed by the Institutional Review Board at Connecticut Children’s and deemed minimal risk. Registered at Clinicaltrials.gov NCT04838834.

### SARS-CoV-2 Spike protein binding domain (S-RBD) IgG antibody responses

To measure SARS-CoV-2 S-RBD IgG antibodies we used a fluorescent bead-based immunoassay developed as previously described (15). We screened patient serum samples for SARS-CoV-2 wild type (WT) S-RBD or K417N, E484K, N501 mutant (mt S-RBD) Spike protein receptor binding domain specific IgG antibodies. Both WT S-RBD and mt S-RBD proteins were Biotinylated and purchased from Acro Biosystems. Tenfold serially diluted serum samples were assayed then analyzed by flow cytometry using iQue Screener Plus (IntelliCyt, MI)(15). Flow cytometry data were analyzed using FlowJo (BD biosciences). Titration curves for each sample were used to normalize the area under the curve (AUC) values to quantitate the antibody levels. Statistical analyses were performed using GraphPad Prism 8.0 software (GraphPad Software).

### Pseudotyped virus neutralization assay

We developed a sensitive SARS-CoV-2 neutralization assay (15) by incorporating WT SARS-CoV-2 Spike protein or with N501Y mutation representing the current prevalent B.1.1.7 (alpha variant) mutation in the US, onto lentiviruses and measuring pseudoviral entry into ACE2 over-expressing 293 cells (293-ACE2). To determine the half-maximal inhibitory concentration (NT50), three-fold serially diluted serum samples from different patients were incubated with RFP-encoding WT SARS-CoV-2 or alpha variant pseudotyped virus at 0.2 multiplicity of infection (MOI) for 1 hour at 37**°**C degrees. The mixture was subsequently incubated with 293-ACE2 cells for 72h hours after which cells were collected, washed with FACS buffer (1xPBS + 2% FBS) and analyzed by flow cytometry using BD FACSymphony A5 analyzer. Percent infection obtained was normalized for samples derived from cells infected with WT SARS-CoV-2 or alpha variant pseudotyped virus in the absence of serum. NT50 was determined using 4-parameter nonlinear regression (GraphPad Prism 8.0).

### Data Analysis and Statistical Methods

To determine patient characteristics and the humoral immune response to SARS-CoV-2, correlations between antibody AUC levels, 50% neutralization titer (NT50) values and demographics of the study subjects were analyzed. All immunological data were entered into a common Excel-based database. These data were then merged with cohort information drawn from the Redcap database, using study identification numbers. The database was updated monthly as the study proceeded and the monthly updates integrated to provide a longitudinal database for analysis. To compare differences across groups, chi-square and Fisher exact tests were used to compare count or categorical data. T-tests and ANOVA were used to compare mean differences and exact p values are reported. In small sample settings, non-parametric rank-sum tests were used to compare continuous measures. Multiple comparison corrections for pairwise comparisons were used as appropriate. All statistical analyses were performed using GraphPad Prism V8 software. Numbers of repeats for each experiment and sample sizes were described in the associated figure legends.

## Results

### Demographic Characteristics and Clinical Presentation

Of 472 patients with IBD treated in our infusion center, 436 (92%) were enrolled (mean age 17 years, range 2-26 years, 58% male). Forty-four/436 (10.1%) demonstrated IgG antibodies specific to RBD part of the SARS-CoV2 Spike protein (S-RBD), which also contains the main neutralization regions, during the period of study. Demographic and clinical characteristics of the 436 participants including the 392 who remained seronegative and the 44 who were positive are shown in Table 1. The group with anti-SARS-CoV-2 positivity was significantly older than the group without S-RBD IgG antibodies (p=0.02). Twenty-four of the 44 reported a positive PCR test, 22 had a household member who tested positive, and 25 reported a household member who experienced symptoms consistent with infection. Duration between most recent positive PCR test and serum sampling was 3.9 weeks (median 2.6, range 1.0-12.6) in seropositive IBD subjects who had documented PCR. For 3/9 asymptomatic patients where PCR+ date was known the average duration between PCR+ and positive serum was 5 weeks. For the other 6 the time of infection was unknown but less than 12 weeks. All adult control patients were PCR+ (mean age 41 years) with a mean duration between +PCR and serum for study of approximately 6 weeks. All non-IBD pediatric controls (mean age 13 years) were PCR+ and had serum drawn during their initial hospital admission for moderate to severe COVID-19.

**Table 1.** Comparison of demographic and treatment factors by seropositivity in the study population.

| Variable | Total (436) | Seronegative (392) | Seropositive (44) | p |
| --- | --- | --- | --- | --- |
| Age (yrs) | Mean 17.0<br>(range 2-26) | 16.9 (2-26) | 18.2 (11-26) | 0.024 |
| Sex, % male | 253, 58.0% | 222, 56.6% | 31, 70.5% | 0.059 |
| Crohn's disease | 308, 70.6% | 274, 69.9% | 34, 77.3% | 0.273 |
| UC/IBD-U | 128, 29.4% | 118, 30.1% | 10, 22.7% | 0.273 |
| Infliximab | 358, 82.1% | 319, 81.4% | 39, 88.6% | 0.161 |
| Infliximab only | 305, 70.0% | 276, 70.4% | 29, 65.9% | 0.549 |
| Infliximab/<br>methotrexate | 53, 12.2% | 43, 11.0% | 10, 22.7% | 0.071 |
| Vedolizumab | 78, 17.9% | 73, 18.6% | 5, 11.4% | 0.301 |
| Vedolizumab only | 74, 17.0% | 69, 17.6% | 5, 11.4% | 0.398 |
| Vedolizumab/<br>methotrexate | 4, 0.9% | 4, 1.1% | 0, 0% | 1.0 |
| Corticosteroids | 15, 3.8% | 15, 3.8% | 0, 0% | 0.382 |

No seropositive participants in the IBD cohort were hospitalized. Nine of 44 (20%) participants were asymptomatic; 3 had a history of positive PCR, one had a family member with a history of COVID. The other 5 were only known to have had SARS-CoV-2 infection by serology. Reported symptoms included headache 27/44 (61%), anosmia/ageusia 18/44 (41%), myalgias 16/44 (36%), ≥3 days of cough 14/44 (32%), anorexia 11/44 (25%), fever ≥101°F 10/44 (23%), sore throat 10/44 (23%), diarrhea 7/44 (16%), vomiting 2/44 (5%). No patient had a flare of their stable IBD following infection with SARS-CoV-2 infection.

Thirty-nine of 358 (10.9%) infliximab treated patients were seropositive compared to 5/78 (6.4%) vedolizumab patients (chi-square, p=0.164). A total of 53 patients were receiving infliximab and methotrexate combination therapy and 10 of these (18.9%) were found to be antibody positive. No patients on vedolizumab plus methotrexate (n=4) were found to be positive. No patient who was receiving corticosteroids at the time of serum sampling proved seropositive (n=15). No patients were receiving thiopurines.

### Magnitude of serological response

We first compared anti-S-RBD IgG responses in our IBD study population to non-IBD adults and pediatric patients with COVID-19. Serum anti-S-RBD IgG was significantly lower than either comparator group, most notably (9.9x difference) between IBD patients and non-IBD adults (Figure 1a). There was no significant difference between the biologic treatment groups. We next assessed the neutralizing activity of the serum from each of the 3 study groups using a pseudovirus test to block cell entry, as described (15). While the neutralization titer (NT50) between adult and pediatric non-IBD groups were similar, both were significantly higher compared to the IBD group (Figure 1b). No neutralizing activity was observed in 5/44 of the IBD group as well as 1/23 of the adult non-IBD subjects and 1/11 of the pediatric non-IBD subjects. We did not find any significant differences between the biologic treatment groups. There was also significant correlation between NT50 and S-RBD IgG levels (Supplemental Figure 1a)

**Figure 1:**
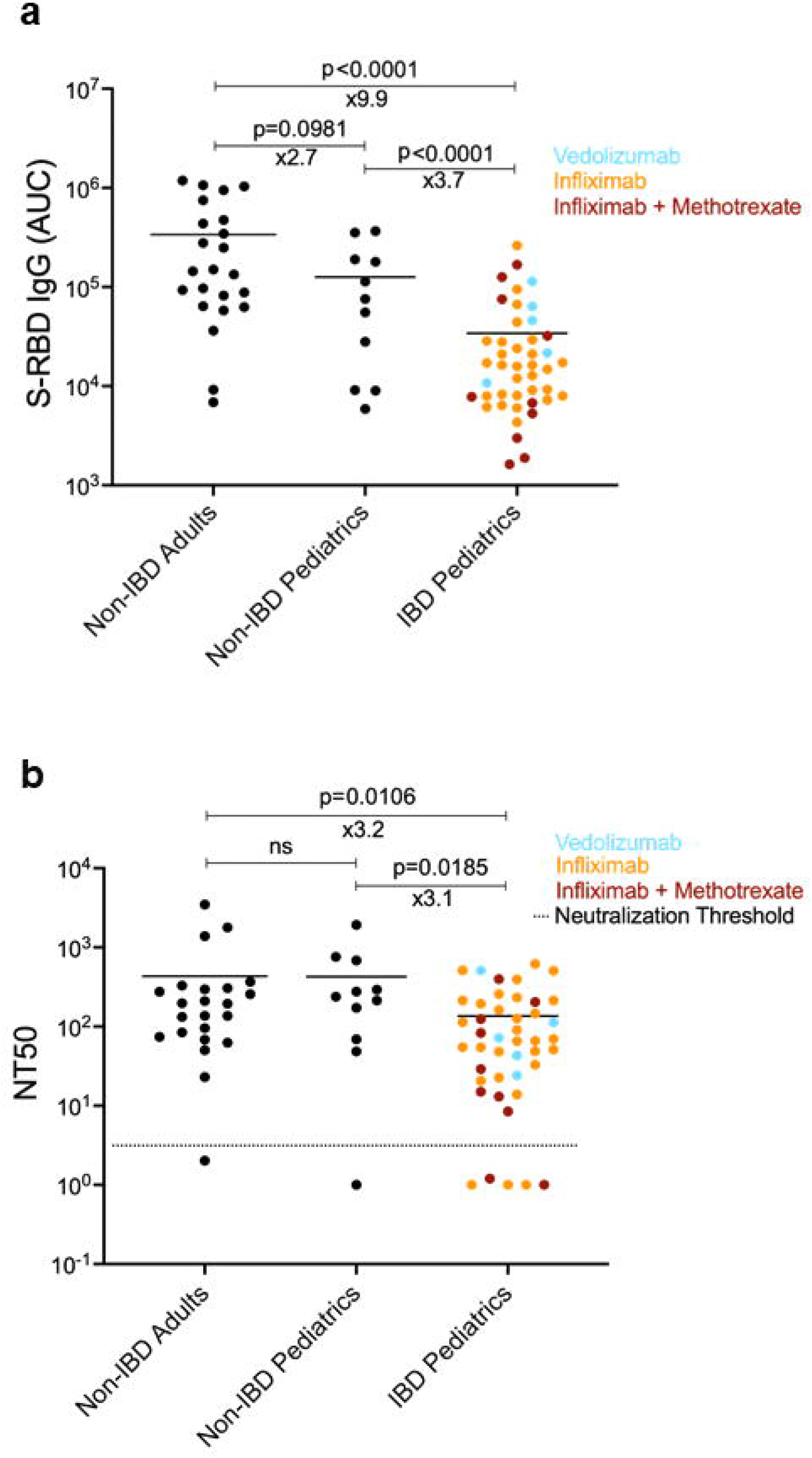
Comparison of the anti-spike protein IgG antibody levels and neutralization titers of adult non-IBD (Inflammatory Bowel Disease), non-IBD pediatric and IBD pediatric subjects receiving biologic therapy. **a**. SARS-CoV-2 Spike protein receptor binding domain (S-RBD) specific IgG antibody levels measured in serum of non-IBD adult, non-IBD pediatric and IBD pediatric subjects. Area under the curve (AUC) values of serum antibodies were calculated from reciprocal dilution curves in antibody detection assay as described in the Methods (n=23 for non-IBD adults, n=11 for non-IBD pediatrics and n=44 for IBD pediatrics). Blue, orange, and red dots indicate the subjects with Vedolizumab monotherapy, Infliximab monotherapy, and Infliximab + Methotrexate co-therapy, respectively. Horizontal bars show the mean value. x values under the significance bars represent the fold changes between the mean values of groups. **b**. Half-maximal neutralization titer (NT50) values of adult non-IBD, pediatric non-IBD and pediatric IBD subjects (n=23 for non-IBD adults, n=11 for non-IBD pediatrics and n=44 for IBD pediatrics). NT50s of serum samples were measured via a neutralization assay using SARS-CoV-2 pseudotyped lentiviruses as described in the Methods. Dotted lines indicate the neutralization threshold which was NT50 of 5. Two-tailed Mann-Whitney U test was used to determine the statistical significances.

### Durability of antibody response to SARS-CoV-2 Spike protein

We next asked to what extent the SARS-CoV-2 S-RBD IgG response to infection in the adult non-IBD subjects compared to the IBD patients was sustained over a period of 6 months. Repeated sampling was performed in the adult subjects at approximately 2-month intervals and in the IBD subjects at approximately 3 and 6 months following initial positive sample. As expected, S-RBD specific IgG decreased over time in the adult cohort (Figure 2a) and in IBD subjects (Figure 2b); however, the magnitude of decrement between baseline and 6-months later appeared to be higher in IBD patients (3.7x decline). Interestingly, the ability to neutralize the virus was retained in almost all adult subjects after 2 months without significant further decrease (Figure 2c). However, neutralization capacity markedly decreased over time and by 6 months (10.6x decline) the majority of the IBD patients had lost their neutralizing activity (Figure 2d). Specific biologic regimen for the IBD patients did not show statistical significance.

**Figure 2:**
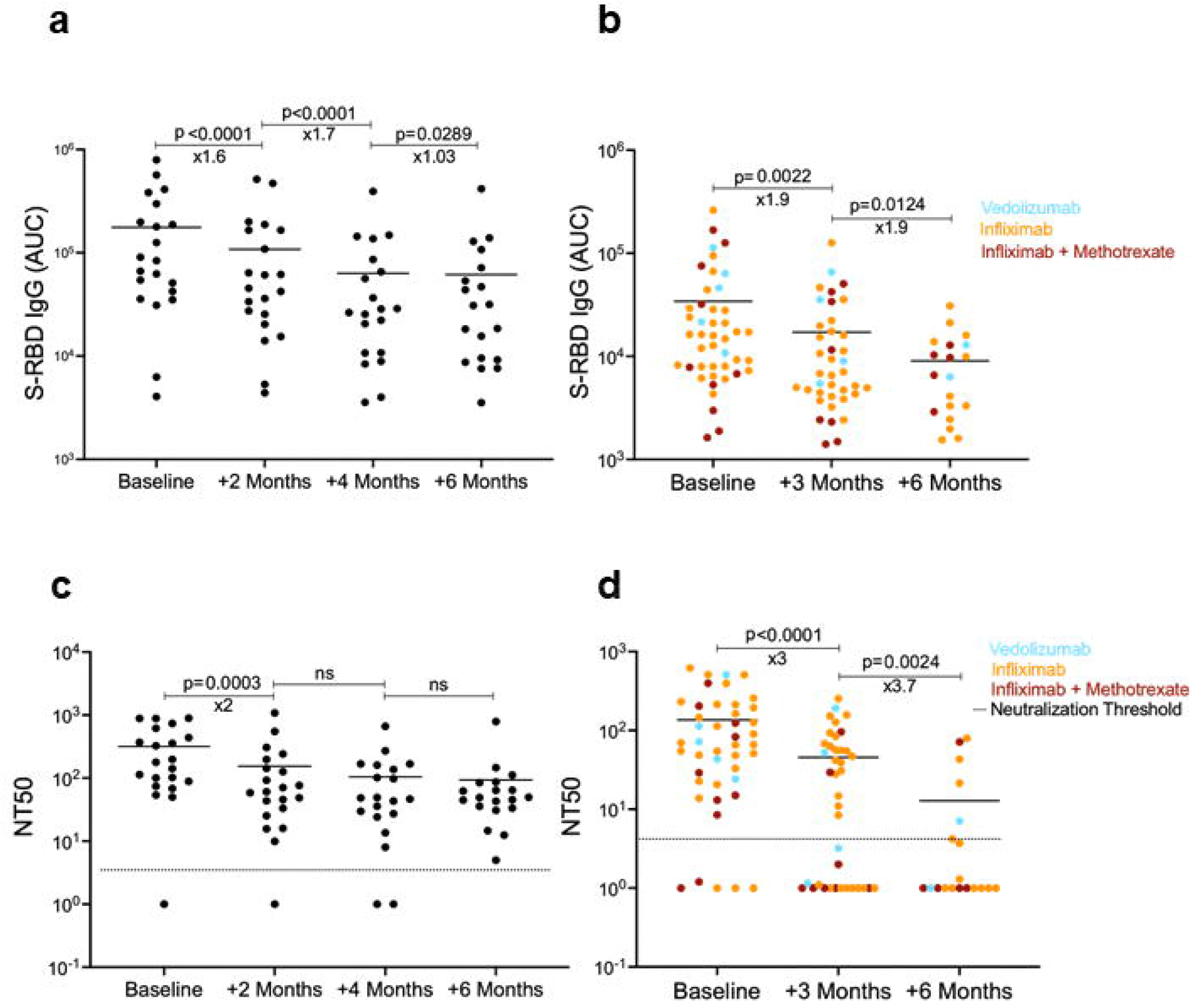
Antibody levels and neutralization titers of non-IBD adults and IBD pediatric subjects at different time points. **a**. S-RBD IgG levels of non-IBD adults at the time of diagnosis and 2, 4, 6 months thereafter. (n= 21 for baseline and +2 months, n=20 for +4 months, n=19 for +6 months) **b**. S-RBD IgG levels of Pediatric IBD subjects at the time of diagnosis and 3 and 6 months thereafter. (n= 44 for baseline, n=39 for +3 months, n=19 for +6 months). **c**. Neutralization titer (NT50) of COVID-19 infected non-IBD adults and of **d**. IBD subjects over 6-month period. Horizontal bars show the mean value. Blue, orange, and red dots indicate the subjects with Vedolizumab monotherapy, Infliximab monotherapy, and Infliximab + Methotrexate co-therapy, respectively. x values under the significance bars represent the fold changes between the mean values of groups. Dotted lines indicate the neutralization threshold which was NT50 of 5. Two-tailed Mann-Whitney U test was used to determine the statistical significance.

### SARS-CoV-2 Spike specific antibody response following vaccination

We compared the anti-S-RBD antibody response to natural infection versus the response to immunization (n= 33 IBD subjects). 28 subjects received a mRNA vaccine (either Pfizer-BioNTech (n=21) or Moderna (n=7)) and 5 the adenovirus vector vaccine (AVV) (Johnson & Johnson). Four out of 33 vaccinated IBD patients had documented previous SARS-CoV-2 infection. Serum sampling post second dose vaccination for the mRNA vaccine was obtained at a mean of 3.3 weeks, range 1 to 10 weeks. For the 5 subjects receiving the adenovirus vector vaccine the duration between vaccination and serology had a mean of 3.1 weeks, range 1.6 to 3.6 weeks. S-RBD IgG responses were significantly higher (∼15x) following vaccination in comparison to natural infection in the IBD subgroup (Figure 3a). Moreover, all IBD subjects had higher neutralizing IgG post vaccination compared to natural infection (∼10x), including patients receiving both infliximab monotherapy and infliximab along with methotrexate (Figure 3b). Further, NT50 was highly correlated with S-RBD IgG levels in vaccinated individuals (Supplemental Figure 1b)

**Figure 3:**
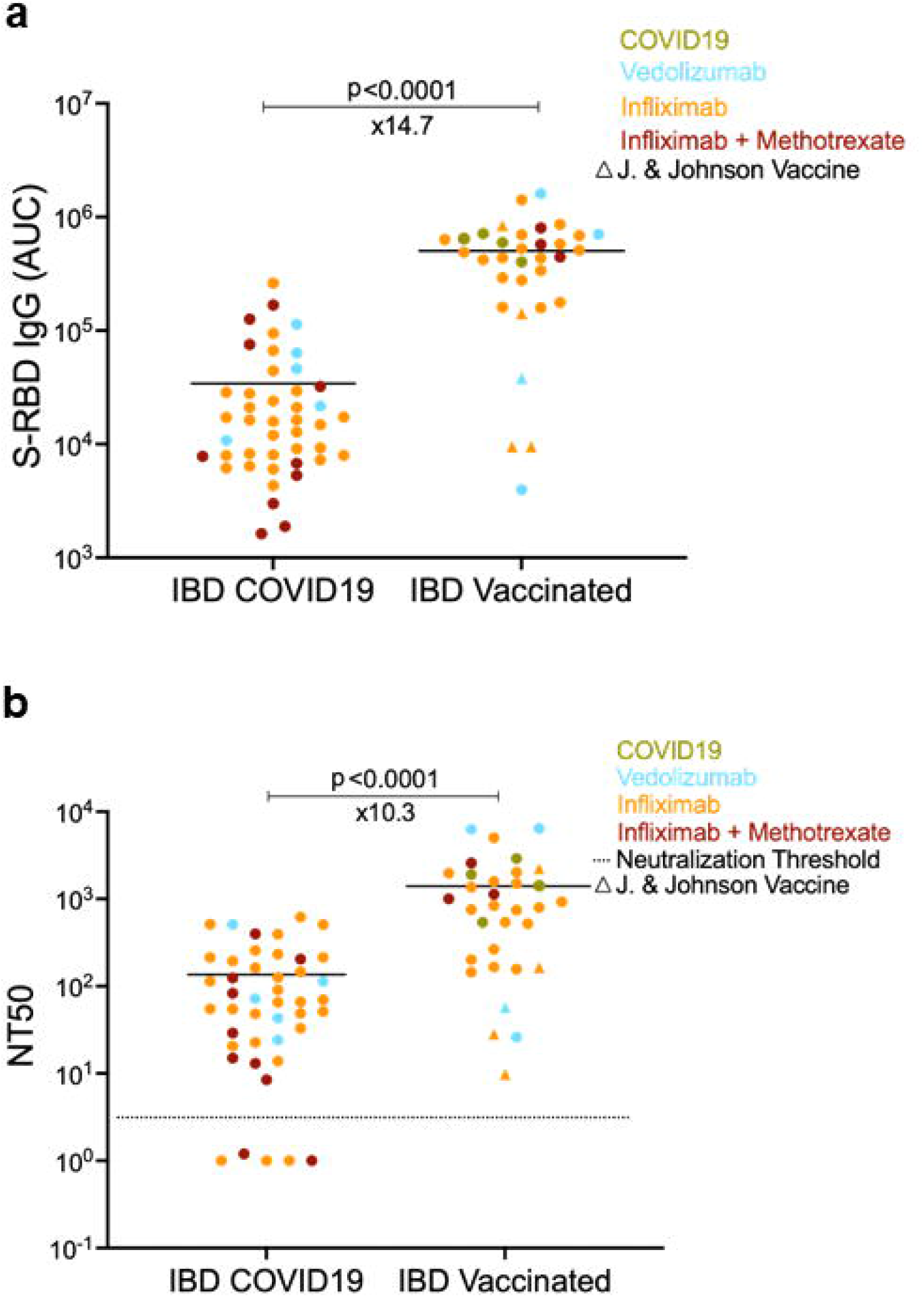
Comparing the antibody levels and neutralization titers of COVID-19 seropositive and vaccinated IBD pediatric subjects. **a**. S-RBD IgG levels of COVID-19 seropositive pediatric IBD subjects (n=44) and vaccinated pediatric IBD subjects (n=33), respectively. **b**. Neutralization titers (NT50) post COVID19 and vaccination in IBD subjects. The average durations were 4.0 weeks after the first PCR+ test for IBD COVID-19 subjects and 3.2 weeks after the fully vaccination date for IBD vaccinated subjects. Blue, orange, and red dots indicate the subjects with Vedolizumab monotherapy, Infliximab monotherapy, and Infliximab + Methotrexate co-therapy, respectively. Green dots represent the COVID-19 seropositive subjects among the vaccinated group. Circles indicate RNA vaccines with 2 doses in the vaccinated group whereas triangles show Johnson & Johnson adenovirus vaccine which was administered once. Horizontal bars show the mean value. x values under the significance bars represent the fold changes between the mean values of groups. Dotted lines indicate the neutralization threshold which was NT50 of 5. Two-tailed Mann-Whitney U test was used to determine the statistical significance.

We next compared anti-S-RBD IgG responses in our IBD cohort to wild-type and a mutated SARS-CoV-2 spike protein. In response to natural infection, mean S-RBD IgG levels were 3.7x lower to the mutant spike protein when compared to the wild-type spike protein (p< 0.0001) (Figure 4a). Whereas we did not see a significant difference between WT and mutant strain spike protein S-RBD-IgG levels in response to vaccination (Figure 4b). Indeed, S-RBD IgG responses to the mutant spike protein were 34.3x lower in response to natural infection when compared to vaccine induced antibodies (Figure 4c). The neutralization of pseudovirus expressing wild-type spike protein vs with N501Y mutation (main mutation in B.1.1.7 or alpha variant) was very similar post vaccination (Supplemental Figure 2a) and highly correlated with the S-RBD antibody levels (Supplemental Figure 2b). Taken together, these findings suggest that the vaccination of children and young adults with IBD elicits much higher antibody responses capable of overcoming immune evading SARS-CoV-2 mutations.

**Figure 4:**
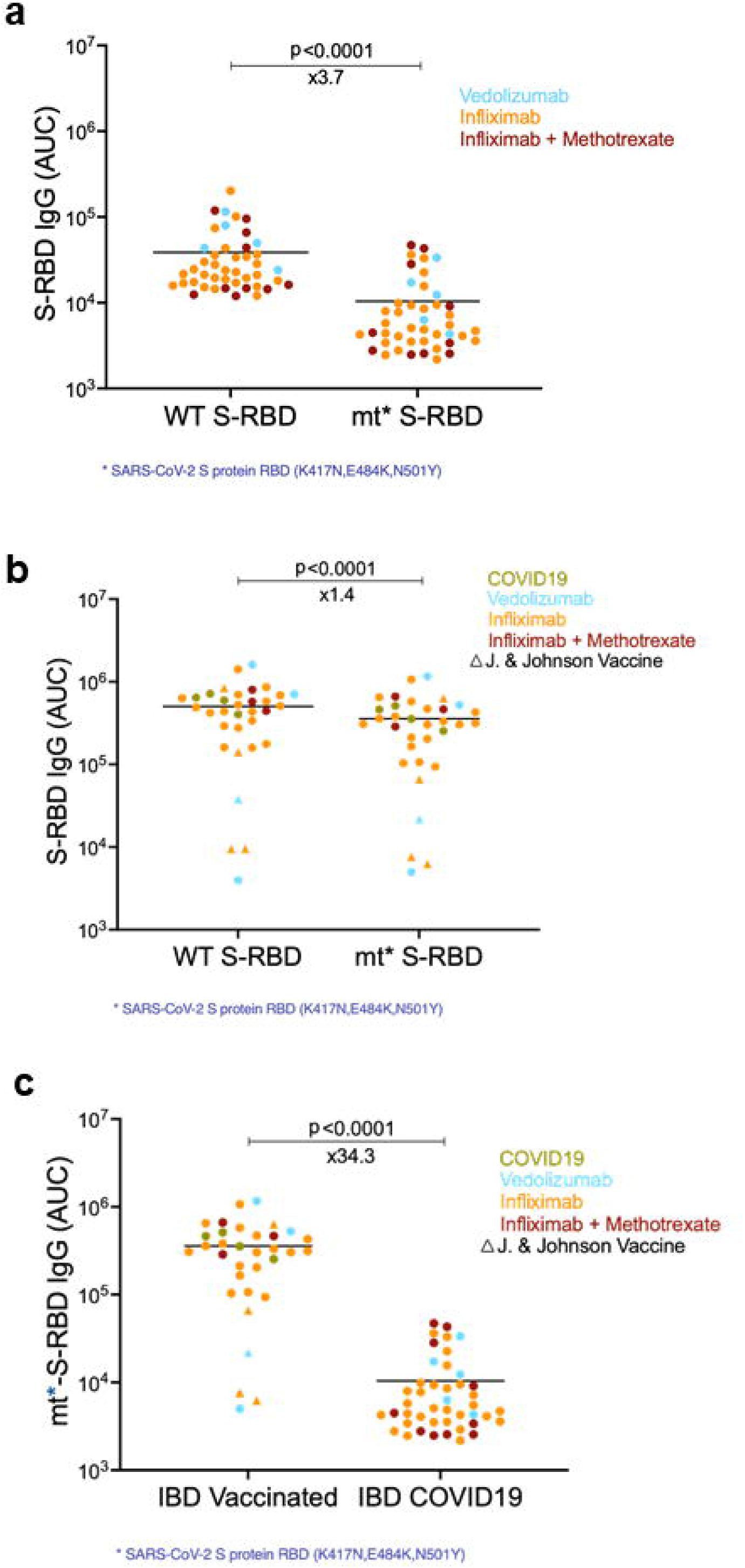
Specific antibody levels of wild-type S-RBD and mutant S-RBD in the serum of COVID-19 seropositive and vaccinated IBD pediatric subjects. **a** Levels of anti-spike protein IgG antibodies to wild-type (WT S-RBD) and mutant S-RBD (mt* S-RBD) in COVID-19 seropositive (n=44) IBD serum samples, and **b**. in vaccinated (n=33) IBD subjects. respectively. Mutations in S-RBD protein were K417N, E484K and N501Y. Blue, orange, and red dots indicate the subjects with Vedolizumab monotherapy, Infliximab monotherapy, and Infliximab + Methotrexate co-therapy, respectively. Green dots represent the COVID-19 seropositive subjects among the vaccinated group. Circles indicate RNA vaccines with 2 doses in the IBD vaccinated group whereas triangles show Johnson & Johnson adenovirus vaccine which was administered once. **c**. Comparison of the levels of mutant S-RBD specific IgG antibodies in vaccinated and COVID-19 seropositive IBD pediatrics serum samples. Horizontal bars show the mean value. x values under the significance bars represent the fold changes between the mean values of groups. Two-tailed Wilcoxon test was used to determine the statistical significance in **a, b** and Mann-Whitney U tests was used for the statistical calculation in **c**.

## Discussion

Our data have significant and immediate implications for patients with IBD receiving biologic therapy and potentially for others with chronic inflammatory disease receiving similar therapies. The decreased S-RBD IgG response compared to adult non-IBD controls and the relatively rapid disappearance of neutralizing antibodies post-infection implies increased risk for reinfection. However, the strong response to vaccination in these subjects with the generation of S-RBD neutralizing antibody of higher titer than natural infection, underscores the critical importance of immunization in this patient group.

During the period we performed our antibody screens SARS-CoV-2 infection in the United States (16) was around 10% underscoring that our study population, which also had around 10% evidence of SARS-CoV-2 infection, was not at increased risk of infection. The clinical expression of disease was mild as has been observed (1, 2). Yet, our data differ from the study in the United Kingdom (13) where less than half of PCR+ patients with IBD receiving infliximab were found to have antibodies to SARS-CoV-2. In our sampling, we found 44/436 (about 10%) to have S-RBD antibodies, including 24 IBD patients who had tested PCR+ and 23/24 had S-RBD IgG antibodies and had detectable neutralizing antibody. Our overall prevalence of antibody positivity to SARS-CoV-2 was about twice that noted in the United Kingdom study, likely reflecting the epidemiology of infection in Connecticut compared to the United Kingdom.

Data are now emerging on the response to immunization with mRNA and other vaccines. A recent study demonstrated that infliximab was associated with attenuated immunogenicity to a single dose of the BNT162b2 and ChAdOx1 nCoV-19 SARS-CoV-2 vaccines (17). However, vaccination after SARS-CoV-2 infection, or a second dose of vaccine, led to seroconversion in most patients. Our data, acquired after 2 doses of mRNA vaccine showed robust seroconversion despite infliximab therapy and all patients had neutralizing antibody. We did not sample patients after a single dose. Our observations strongly support the recent vaccination endorsement by the International Organization for the Study of IBD (18) and lessen theoretical concerns about diminished response to immunization for influenza (19, 20) and hepatitis B (21) as has been reported in patients on anti-TNF therapy. Indeed, a recent systematic review and meta-analysis suggested that immunosuppressive therapy does not significantly reduce immunogenicity of vaccinations in children with IBD (22).

The potential effect of anti-metabolites on response to immunization remains unclear and a recent report in solid organ transplant patients suggested a lessened response to mRNA vaccination (23). In adult patients with rheumatoid arthritis a temporary discontinuation of methotrexate for 2 weeks prior to influenza vaccination improved immunogenicity (24). Methotrexate is often used either alone or in combination with anti-TNF agents in the treatment of IBD. In our cohort, the number of patients on combination infliximab and methotrexate who had also been vaccinated was too small to compare, albeit all had similar S-RBD IgG levels. Future studies assessing larger numbers of IBD patients on combination infliximab and anti-metabolites would be useful to determine if the addition of anti-metabolite therapies lessen the antibody responses to either natural infection or immunization. Finally, 4 of our immunized patients were documented to have had SARS-CoV-2 infection prior to immunization and all received 2 doses of a mRNA vaccine. It has been suggested that only one dose may be necessary in healthy subjects with prior infection (25) but we did not obtain samples after one dose of vaccine in our patient cohort, thus the impact of the second dose remains to be determined.

Our study has several significant strengths. We performed longitudinal surveillance on a defined population allowing us to identify both symptomatic and asymptomatic infection. In contrast to previous studies, which just measured antibody levels, we determined neutralization activity of the anti-S-RBD IgG to both wild-type and variant virus. Thus, we were able to determine in an assay assumed to be a surrogate for protection from clinical infection, that most of our patients generated titers of neutralizing antibody post-infection that rapidly declined over a 6-month period. There were also several limitations. We did not have control data from IBD patients not receiving biologics, particularly those on methotrexate or thiopurine monotherapy. We also do not yet have longitudinal data on the durability of antibody response following vaccination. Finally, we did not test antibody neutralization ability from our patients against emerging SARS CoV-2 variants which may reveal lower titers and a more rapid decline in protective levels. However, given almost perfect correlation of S-RBD IgG levels with the NT50 (Supplemental Figure 1b) and a 34.3x lower antibody response to mutant S-RBD in natural infection compared to vaccine (Figure 4c), we anticipate neutralization activity to decline as we observed for the S-RBD antibody level.

In conclusion, the humoral response to SARS-CoV-2 infection in children and young adults was less than in adult and pediatric non-IBD controls and neutralizing antibody was absent in most infected IBD patients by 6 months. The robust response to immunization was reassuring and supports the utility of vaccinating these patients. Further data will be required to understand the durability of the response to the vaccine, whether previous infection will enhance the antibody response to the vaccine in this patient group lessening the need for a second dose, and whether chronic anti-metabolite administration will have a mitigating influence over time.

## Data Availability

Available upon request

## Acknowledgements

The authors thank the patients and families cared for at the Connecticut Children’s Infusion Center for their participation, the nursing team at the Connecticut Children’s Infusion Center for their strong support of this study, Kristen Volz for logistical support, and the Mandell Braunstein IBD Research Chair Fund (JH), the David and Geri Epstein Foundation Fund (JH), the Ellen Oland Endowed Fund for Pediatric IBD Research (JH) and the Achelis and Bodman Foundation (DU) and NIH grant U19 AI142733-01 (DU) for financial support.

## Author contributions

Dr. Hyams had full access to all the data in the study and takes responsibility for the integrity of the data and accuracy of the data analysis.

Concept and design: Hyams, Dailey, Kozhaya, Dogan, Hopkins, Brimacombe, Schreiber, Salazar, Unutmaz.

Acquisition, analysis, and interpretation of the data: Hyams, Dailey, Hopkins, Kozhaya, Dogan, Liang, Lapin, Herbst, Grandonico, Karabacak, Brimacombe, Schreiber, Salazar, Unutmaz

Drafting of the manuscript: Hyams, Dailey, Hopkins, Kozhaya, Dogan, Brimacombe, Salazar, Unutmaz

Critical revision of the manuscript for important intellectual content: Hyams, Dailey, Hopkins, Kozhaya, Dogan, Lapin, Liang, Herbst, Brimacombe, Grandonico, Karabacik, Schreiber, Salazar, Unutmaz

Supervision: Hyams, Salazar, Unutmaz

## SUPPLEMENTAL FIGURE LEGENDS

**Supplemental Figure 1: Correlation of neutralization titers and wild-type S-RBD specific antibody levels in COVID-19 seropositive and vaccinated pediatric IBD subjects**.

**a**. S-RBD specific IgG antibody AUC levels correlated with. NT50 in COVID-19 seropositive (n=44), and **b**. vaccinated IBD (n=33) subjects in serum. Blue, orange, and red dots indicate the subjects with Vedolizumab monotherapy, Infliximab monotherapy, and Infliximab + Methotrexate co-therapy, respectively. Green dots represent the COVID-19 seropositive subjects among the vaccinated group. Circles indicate RNA vaccines with 2 doses in the IBD vaccinated group whereas triangles show Johnson & Johnson adenovirus vaccine which was administered once. Dotted lines indicate the neutralization threshold which was NT50 of 5. Spearman’s test was used to determine the statistical significance.

**Supplemental Figure 2: Neutralization capacity in vaccinated pediatric IBD subjects’ serum against wild-type and N501Y-mutation variant SARS-CoV-2 Spike protein pseudotyped lentiviruses**.

**a**. Comparison of neutralization titers of vaccinated IBD pediatric subjects’ serum against wild-type and N501Y Spike protein pseudotyped lentiviruses (n=33). NT50 values were measured with lentiviruses pseudotyped with wild-type and N501Y-mutated SARS-CoV-2 spike proteins as described in the Methods. Blue, orange, and red dots indicate the subjects with Vedolizumab monotherapy, Infliximab monotherapy, and Infliximab + Methotrexate co-therapy, respectively. Green dots represent the COVID-19 seropositive subjects among the vaccinated group. Circles indicate RNA vaccines with 2 doses whereas triangles show Johnson & Johnson adenovirus vaccine which was administered once. Dotted lines indicate the neutralization threshold which was NT50 of 5. Two-tailed Wilcoxon test was used to determine the statistical significance. **b**. Correlation of NT50 values of vaccinated IBD subjects (n=33) measured using wild-type and N501Y-mutated Spike protein pseudotyped lentiviruses. Two-tailed Spearman’s was used to determine the statistical significance.

## References

1. Brenner EJ, Ungaro RC, Gearry RB, Kaplan GG, Kissous-Hunt M, Lewis JD, et al. Corticosteroids, But Not TNF Antagonists, Are Associated With Adverse COVID-19 Outcomes in Patients With Inflammatory Bowel Diseases: Results From an International Registry. Gastroenterology. 2020;159(2):481–91 e3.

2. Brenner EJ, Pigneur B, Focht G, Zhang X, Ungaro RC, Colombel JF, et al. Benign Evolution of SARS-Cov2 Infections in Children With Inflammatory Bowel Disease: Results From Two International Databases. Clin Gastroenterol Hepatol. 2021;19(2):394–6 e5.

3. Allocca M, Fiorino G, Zallot C, Furfaro F, Gilardi D, Radice S, et al. Incidence and Patterns of COVID-19 Among Inflammatory Bowel Disease Patients From the Nancy and Milan Cohorts. Clin Gastroenterol Hepatol. 2020;18(9):2134–5.

4. Ungaro RC, Brenner EJ, Gearry RB, Kaplan GG, Kissous-Hunt M, Lewis JD, et al. Effect of IBD medications on COVID-19 outcomes: results from an international registry. Gut. 2021;70(4):725–32.

5. Zhou Z, Ren L, Zhang L, Zhong J, Xiao Y, Jia Z, et al. Heightened Innate Immune Responses in the Respiratory Tract of COVID-19 Patients. Cell host & microbe. 2020;27(6):883–90 e2.

6. Vardhana SA, Wolchok JD. The many faces of the anti-COVID immune response. J Exp Med. 2020;217(6).

7. McKechnie JL, Blish CA. The Innate Immune System: Fighting on the Front Lines or Fanning the Flames of COVID-19? Cell host & microbe. 2020;27(6):863–9.

8. Mathew D, Giles JR, Baxter AE, Greenplate AR, Wu JE, Alanio C, et al. Deep immune profiling of COVID-19 patients reveals patient heterogeneity and distinct immunotypes with implications for therapeutic interventions. bioRxiv. 2020.

9. Huang AT, Garcia-Carreras B, Hitchings MDT, Yang B, Katzelnick LC, Rattigan SM, et al. A systematic review of antibody mediated immunity to coronaviruses: antibody kinetics, correlates of protection, and association of antibody responses with severity of disease. medRxiv. 2020.

10. Dittadi R, Afshar H, Carraro P. The early antibody response to SARS-Cov-2 infection. Clin Chem Lab Med. 2020;58(10):e201–e3.

11. Robbiani DF, Gaebler C, Muecksch F, Lorenzi JCC, Wang Z, Cho A, et al. Convergent antibody responses to SARS-CoV-2 in convalescent individuals. Nature. 2020;584(7821):437–42.

12. Catanzaro M, Fagiani F, Racchi M, Corsini E, Govoni S, Lanni C. Immune response in COVID-19: addressing a pharmacological challenge by targeting pathways triggered by SARS-CoV-2. Signal Transduct Target Ther. 2020;5(1):84.

13. Kennedy NA, Goodhand JR, Bewshea C, Nice R, Chee D, Lin S, et al. Anti-SARS-CoV-2 antibody responses are attenuated in patients with IBD treated with infliximab. Gut. 2021;70(5):865–75.

14. Harris PA, Taylor R, Thielke R, Payne J, Gonzalez N, Conde JG. Research electronic data capture (REDCap)--a metadata-driven methodology and workflow process for providing translational research informatics support. J Biomed Inform. 2009;42(2):377–81.

15. Dogan M, Kozhaya L, Placek L, Gunter C, Yigit M, Hardy R, et al. SARS-CoV-2 specific antibody and neutralization assays reveal the wide range of the humoral immune response to virus. Commun Biol. 2021;4(1):129.

16. Bajema KL, Wiegand RE, Cuffe K, Patel SV, Iachan R, Lim T, et al. Estimated SARS-CoV-2 Seroprevalence in the US as of September 2020. JAMA Intern Med. 2021;181(4):450–60.

17. Kennedy NA, Lin S, Goodhand JR, Chanchlani N, Hamilton B, Bewshea C, et al. Infliximab is associated with attenuated immunogenicity to BNT162b2 and ChAdOx1 nCoV-19 SARS-CoV-2 vaccines in patients with IBD. Gut. 2021.

18. Siegel CA, Melmed GY, McGovern DP, Rai V, Krammer F, Rubin DT, et al. SARS-CoV-2 vaccination for patients with inflammatory bowel diseases: recommendations from an international consensus meeting. Gut. 2021;70(4):635–40.

19. Andrisani G, Frasca D, Romero M, Armuzzi A, Felice C, Marzo M, et al. Immune response to influenza A/H1N1 vaccine in inflammatory bowel disease patients treated with anti TNF-alpha agents: effects of combined therapy with immunosuppressants. J Crohns Colitis. 2013;7(4):301–7.

20. deBruyn J, Fonseca K, Ghosh S, Panaccione R, Gasia MF, Ueno A, et al. Immunogenicity of Influenza Vaccine for Patients with Inflammatory Bowel Disease on Maintenance Infliximab Therapy: A Randomized Trial. Inflamm Bowel Dis. 2016;22(3):638–47.

21. Pratt PK, Jr., David N, Weber HC, Little FF, Kourkoumpetis T, Patts GJ, et al. Antibody Response to Hepatitis B Virus Vaccine is Impaired in Patients With Inflammatory Bowel Disease on Infliximab Therapy. Inflamm Bowel Dis. 2018;24(2):380–6.

22. Dembinski L, Dziekiewicz M, Banaszkiewicz A. Immune Response to Vaccination in Children and Young People With Inflammatory Bowel Disease: A Systematic Review and Meta-analysis. J Pediatr Gastroenterol Nutr. 2020;71(4):423–32.

23. Boyarsky BJ, Ruddy JA, Connolly CM, Ou MT, Werbel WA, Garonzik-Wang JM, et al. Antibody response to a single dose of SARS-CoV-2 mRNA vaccine in patients with rheumatic and musculoskeletal diseases. Ann Rheum Dis. 2021.

24. Park JK, Lee YJ, Shin K, Ha YJ, Lee EY, Song YW, et al. Impact of temporary methotrexate discontinuation for 2 weeks on immunogenicity of seasonal influenza vaccination in patients with rheumatoid arthritis: a randomised clinical trial. Ann Rheum Dis. 2018;77(6):898–904.

25. Krammer F, Srivastava K, Alshammary H, Amoako AA, Awawda MH, Beach KF, et al. Antibody Responses in Seropositive Persons after a Single Dose of SARS-CoV-2 mRNA Vaccine. N Engl J Med. 2021;384(14):1372–4.

