## Supplementary figures and images for "Antibody Responses to SARS-CoV-2 after Infection or Vaccination in Children and Young Adults with Inflammatory Bowel Disease"

### Supplemental Figure 1

#### Slide 1
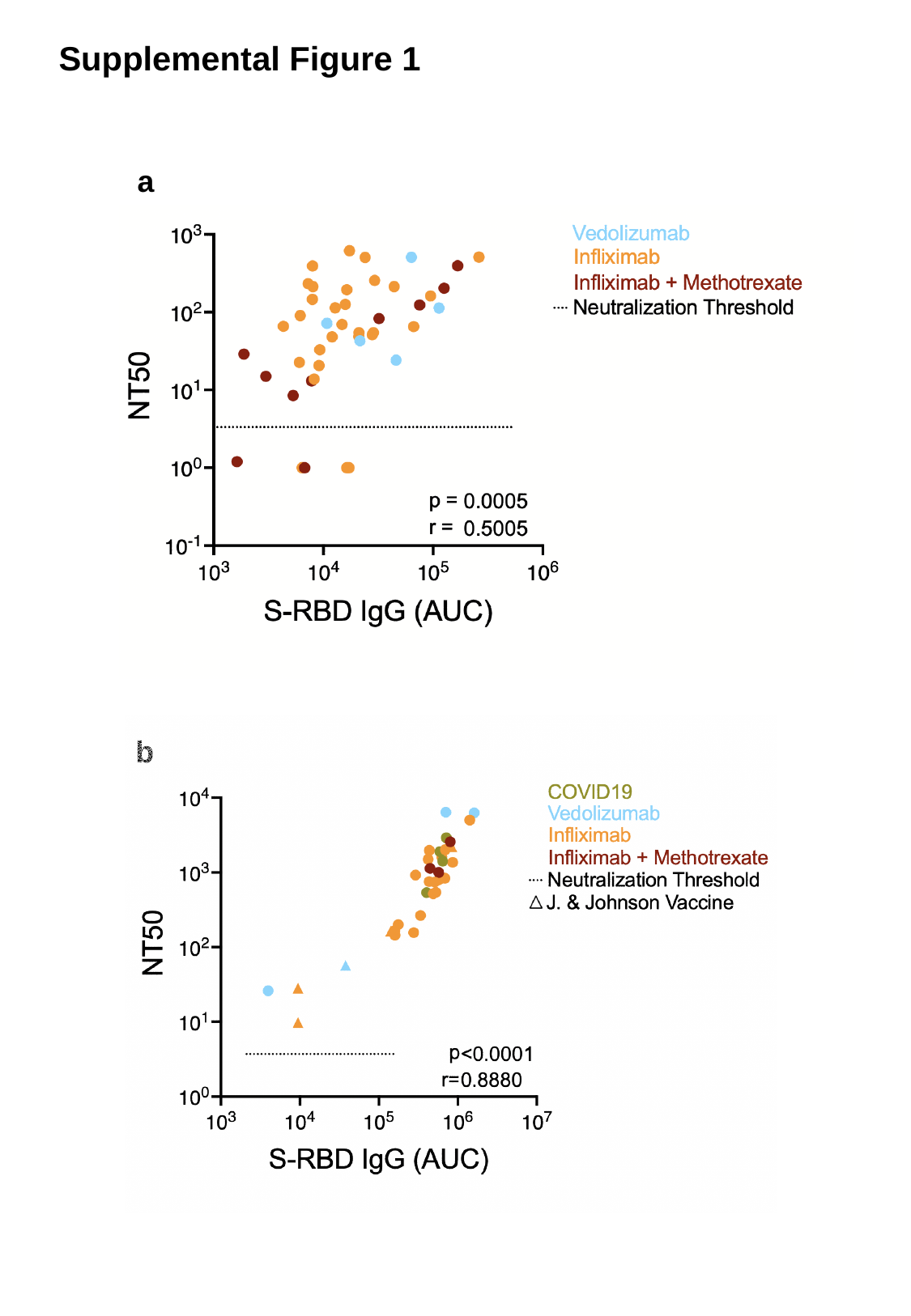

Supplemental Figure 1
a
b

### Supplemental Figure 2

#### Slide 1
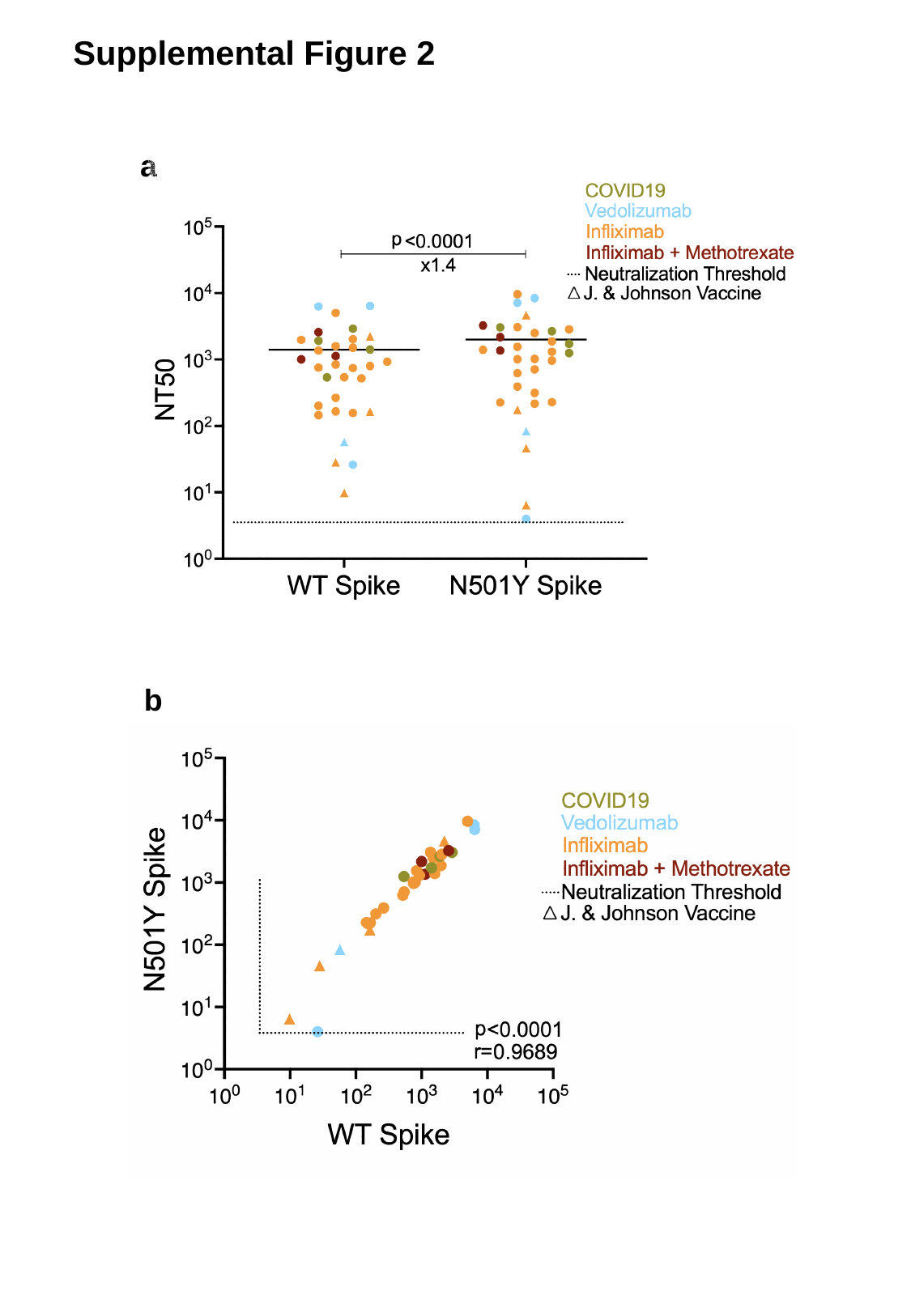

Supplemental Figure 2
a
b
